# Refining clinical algorithms for a neonatal digital platform for low-income countries: a modified Delphi technique

**DOI:** 10.1101/2020.05.23.20111351

**Authors:** Mari Evans, Mark H. Corden, Caroline Crehan, Felicity Fitzgerald, Michelle Heys

## Abstract

**Objectives:** To determine whether a panel of neonatal experts could address evidence gaps in neonatal guidelines by reaching a consensus on four clinical decision algorithms for a neonatal digital platform (NeoTree).

**Design:** Two-round, modified Delphi technique.

**Setting and participants:** Participants were neonatal experts from high-income and low-income countries (LICs).

**Methods:** This was a consensus-generating study. In round one, experts rated items for four clinical algorithms (neonatal sepsis, hypoxic ischaemic encephalopathy, respiratory distress of the newborn, hypothermia) and justified their responses. Items meeting consensus (≥80% agreement) were included. Items not meeting consensus were either excluded, included following revisions or included if they contained core elements of evidence-based guidelines. In round two, experts rated items from round one that did not reach consensus.

**Results:** Fourteen experts participated in round one, ten in round two. Nine were from high-income countries, five from LICs. Experts included physicians and nurse practitioners with an average neonatal experience of 20 years, 12 in LICs. After two rounds, a consensus was reached on 43 of 84 items (52%). Experts consistently stated that items must be in line with local and WHO guidelines (irrespective of the level of supporting evidence or expert opinion). As a result, the final algorithms included 53 items (62%).

**Conclusion:** Four algorithms in a neonatal digital platform were reviewed and refined by consensus expert opinion. Revisions to the NeoTree application were made in response to these findings and will be clinically validated in an imminent study.

**STRENGTHS AND LIMITATIONS OF THIS STUDY:**

➢ In this study, a large number of algorithm items were reviewed and evaluated, and half met consensus for inclusion in the management pathways.
➢ The review was conducted with experts from a broad range of countries and neonatal experience who simultaneously refined the algorithms and highlighted gaps in current evidence, emphasising the need for future research to support international neonatal guidelines.
➢ Our study method meant that experts were not able to meet in person, which might have promoted dialogue that would have allowed greater clarity in their collective opinion.
➢ The representation of neonatal experts from LICs was not as robust as from high-income countries, which may have led to an uneven evaluation of the algorithms.

## INTRODUCTION

Globally, 2.5 million newborns die each year in the first 28 days of life.[1] Most of these deaths (98.5%) occur in low-income countries (LICs), and 40% occur on the first day of life.[2] The neonatal mortality rate has been halved since 1990,[3] but modelling of global newborn mortality data suggests that a further two thirds of current deaths could be prevented if evidence-based solutions were implemented.[2] One of the World Health Organisation (WHO) Sustainable Development Goals (SDG3) is to end preventable deaths of newborns in all countries and to reduce the neonatal mortality rate from the current rate of 18 per 1000 live births to less than 12 per 1000 by 2030.[4] Targeting newborn care in LICs is thus an urgent priority, especially the three most common causes of mortality – infections (36%), prematurity (28%), and intrapartum complications (23%).[2]

Mobile health (m-health) technology and digital platforms are potential approaches to improve the quality of newborn care. They are tools with broad applicability (three quarters of the world now own mobile phones) that are scientifically rigorous and tailored to local patient and healthcare worker (HCW) preferences and resources.[5] A study in Tanzania improved adherence to the Integrated Management of Childhood Illness protocol and assessments of acutely unwell children with an electronic, compared to paper-based, format.[6] M-health and digital platforms can also provide clinical decision algorithms which translate guidelines into context-specific and user-friendly diagnostic aids and management plans. Community HCWs in rural Bangladesh were trained to use a mobile application for neonatal assessment and improved their clinical evaluations, the accuracy of their diagnoses, and their adherence to neonatal management guidelines.[7] Other attempts have been made to introduce health technologies, but most are not available when and where they are needed.[8]

An international team of researchers, clinicians and software developers in the UK, USA, Malawi, Bangladesh and Zimbabwe co-designed and co-developed with Malawian and Zimbabwean HCWs a neonatal digital platform (NeoTree) for facility-based newborn care. It combines immediate digital data capture (which is shared with HCWs via local dashboards), evidence-based algorithmic clinical decision and management support, newborn education and data linkage to national data systems on one platform.[9] The algorithms in the Malawian version of the NeoTree support decisions according to established international[10] and Malawian neonatal guidelines.[11] Local guidelines were combined with international guidelines to accommodate local conditions and to encourage buy-in from local stakeholders. NeoTree is in the phase of application co-development where appropriate revisions to the algorithms are needed to address gaps in evidence for the guidelines.[12] In the absence of extensive trial or epidemiological data, alternative techniques to consolidate best available lower level evidence can be used, such as expert opinion. This study aims to use the modified Delphi technique to determine whether a panel of experts in newborn care can reach a consensus opinion about key clinical decision algorithms used in a digital platform to assist HCWs caring for facility-based unwell newborns in LICs.

## METHODS

### Study design

This study used a two-step modified Delphi technique.[13] The Delphi technique was chosen because it is an effective method of gathering expert knowledge from geographically diverse leaders in the field to address complex clinical problems that lack evidence.

### Recruitment

Twenty-two neonatal experts were invited to participate in the study. This number represented an adequate sample size[14, 15] and permitted a manageable amount of data collection. Participants were recruited if they were a physician or neonatal nurse practitioner with more than ten years neonatal experience (at least three in LICs), neonatal postgraduate training, fluency in English, internet access and willingness to participate. Neonatal experts known to the researchers for their clinical expertise, research and contributions to guideline development in LICs were identified in equal numbers from both LICs and HICs. No financial incentive was offered, but reimbursement for costs of Skype calls was provided for some experts in LICs.

### Algorithms and item generation

The four clinical decision algorithms selected for review were neonatal sepsis, hypoxic ischaemic encephalopathy (HIE), respiratory distress of the newborn and hypothermia. These conditions represent the leading preventable causes of neonatal mortality and are the most difficult to diagnose and manage appropriately in LICs with some of the weakest WHO GRADE recommendations and quality of evidence.[16] For example, the European definition of neonatal sepsis is two or more clinical symptoms and two or more laboratory signs in the presence of, or as a result of, suspected or proven infection.[17] This definition is not possible in LICs where laboratory investigations are not routinely available.

Items were identified by comparing the algorithms side-by-side with the international (WHO) and local neonatal guidelines (Care of the Infant and Newborn in Malawi - COIN) from which they had been derived. This comparison generated a comprehensive list of items where discrepancies in diagnostic parameters and treatment recommendations required expert opinion. Once finalised, the clinical algorithms and list of items (henceforth referred to as questionnaire) were piloted with two paediatricians with neonatal experience in LICs. Ambiguous items were amended accordingly.

### Delphi technique

The questionnaire was circulated by email to the experts with specific instructions at least two weeks before they were interviewed. Each algorithm was verbally and diagrammatically explained with their references specified (i.e., WHO, COIN or NeoTree research team) to aid in decision-making during the interview (online supplementary file 1). Round one interviews were conducted in June and July 2018. Experts were sent up to two reminder emails to schedule their phone or Skype interview. Interviews were conducted privately from a home office. Standardised questions were used to review each item from the questionnaire. Experts were asked to rate their level of agreement for each item using a five-point Likert scale.[18] Each rating was followed by open-ended questions to obtain the experts’ rationale for their response and any amendment or additional items they would propose. All interview data were transcribed using both audio-recordings and notes made during the interview by the facilitator. All responses were anonymised (with participant numbers) and reviewed together with the quantitative results. Based on sample size, the level of agreement for consensus opinion from the panel was set at 80%.[14] Items that met consensus (≥80% agreement) were included or were modified with minor changes to wording based on expert advice. Items that did not meet consensus (<80%) were removed or modified according to the feedback from the expert panel and submitted for the second round. Items that did not meet consensus were still included if they were part of WHO and COIN guidelines so that frontline HCWs continued to follow the current standard of care. A second questionnaire was designed with modified items and expert additions from the first round.

In round two (June and July 2019), this second questionnaire was distributed electronically to the 14 experts from round one (online supplementary file 1). A results summary from round one was sent to the experts, and the full set of anonymised results were made available at their request. Two email reminders were sent to non-responders. Experts again rated items on a Likert scale and explained their ratings. Responses were analysed as described in round one, and items meeting ≥80% consensus were kept for the final NeoTree algorithms.

### Patient and public involvement

While key stakeholders were involved in co-developing the NeoTree digital platform, there was no patient or public involvement in this Delphi study.

### Consent procedures

The goals and processes of the project were explained to the experts in their email invitation, and consent was obtained by email agreement. Experts were verbally informed at the beginning of the first round that their responses would be kept anonymous.

## RESULTS

Twenty-two neonatal experts were invited to participate. Sixteen responded; one declined due to lack of financial incentive, and one declined due to conflict of interest. Demographics of the expert panel are listed in Table 1.

**Table 1.** Characteristics of the Delphi panel from round one

|  | **Characteristics** | **N = 14 (%)** |
| --- | --- | --- |
| Location | Experts from HICs | 9 (64) |
|  | Experts from LICs | 5 (36) |
| Level of expertise | Neonatologist | 6 (43) |
|  | Paediatrician | 6 (43) |
|  | ANNP | 2 (14) |
| Years of neonatal experience post medical school<br>(mean ± SD) | Overall | 20 (± 12) |
| | In LICs | 12 (± 7) |
| Work experience in low-income countries | Africa | 14 (100) |
|  | Asia | 7 (50) |
|  | Central America | 4 (28) |
| Country of medical degree | United Kingdom | 7 |
|  | United States | 2 |
|  | South Africa | 2 |
|  | Zimbabwe | 1 |
|  | Rwanda | 1 |
|  | Sudan | 1 |

ANNP, advanced neonatal nurse practitioner; HICs, high-income countries; LICs, low-income countries

### Round One

Fourteen experts (63% response rate) completed round one. Interviews averaged 73 minutes (40- 110 minutes). Thirty-four items (45%) reached consensus (Figure 1). These items were either: (1) included, unmodified (32%); (2) included, modified (11%); or (3) changed for clarification in the second round (2%). Items that did not reach consensus (55%) were either: (1) excluded from the revised algorithm (30%); (2) included because they were part of WHO/COIN guidelines or the Thompson score (11%);[19] or (3) changed and submitted for a second round (14%) (Table 2).

**Table 2.**
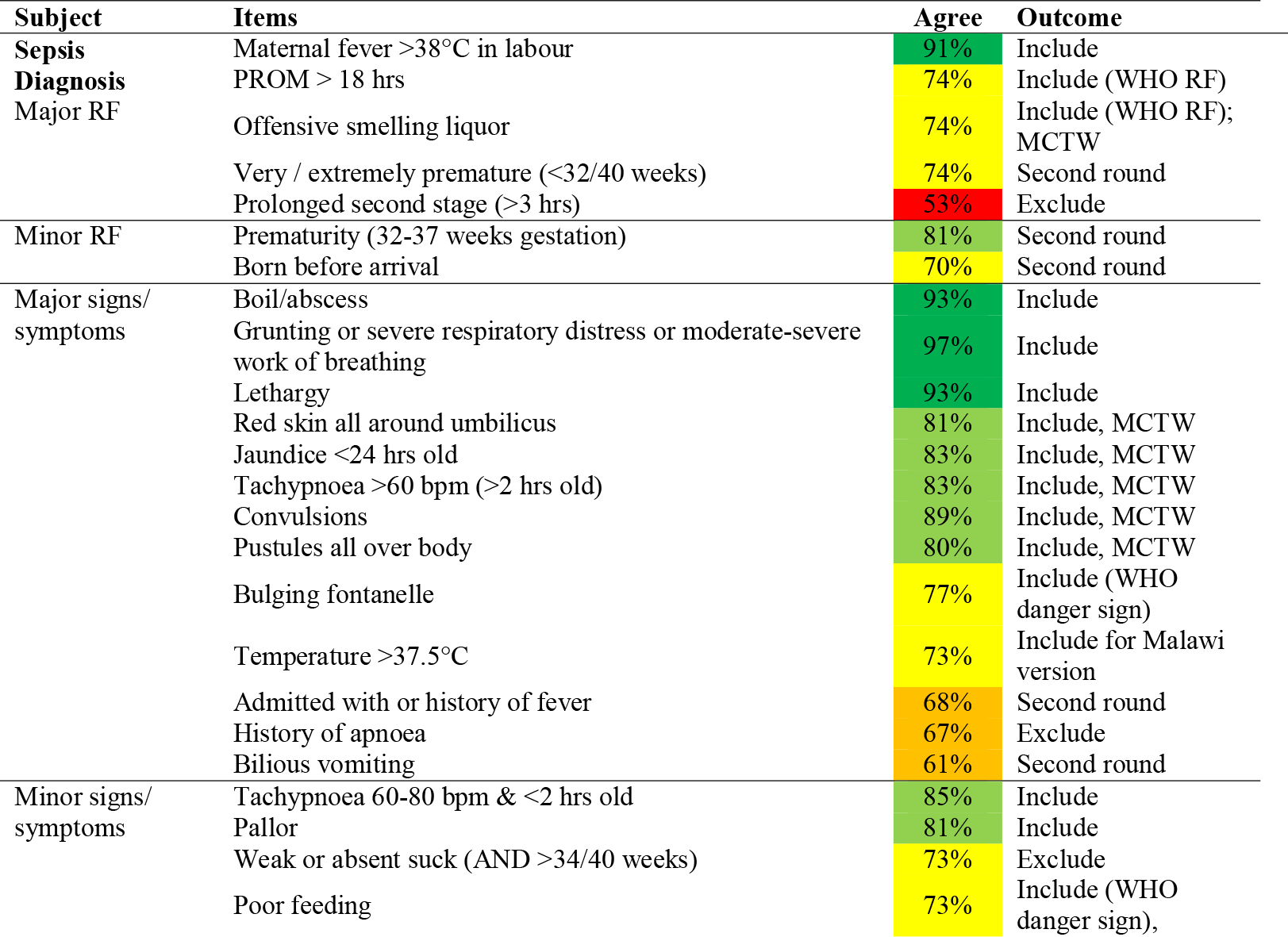
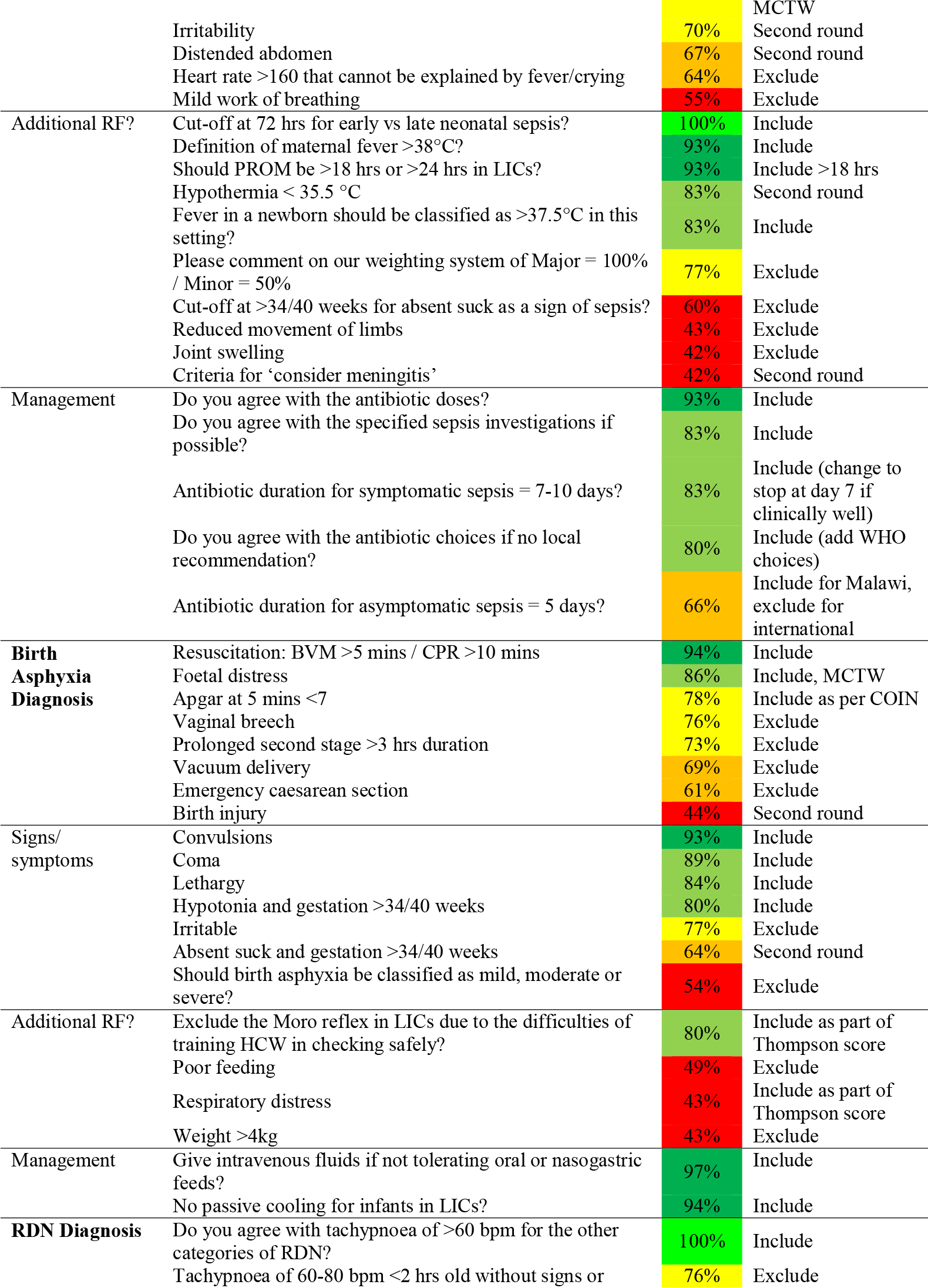
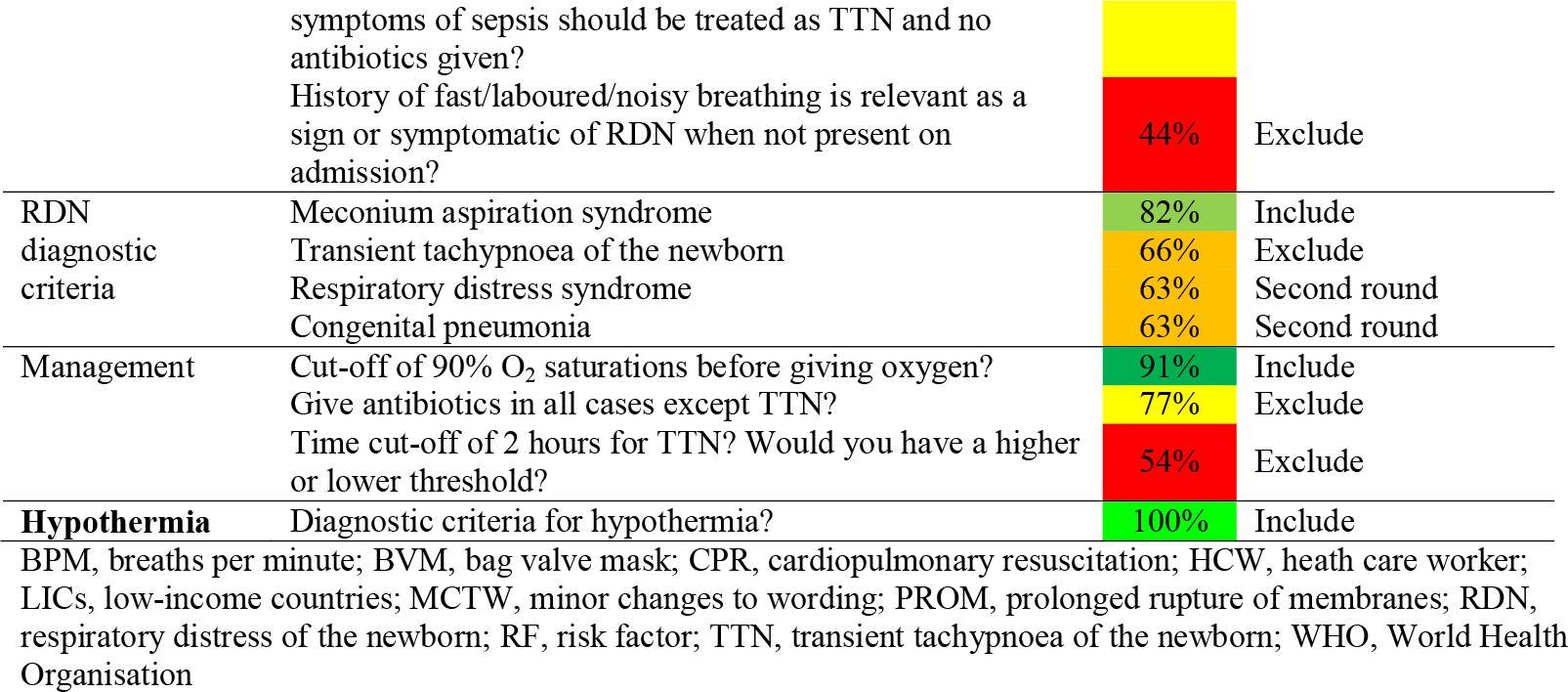
Round one heat chart to show which items met consensus and their outcomes.

**Figure 1.**
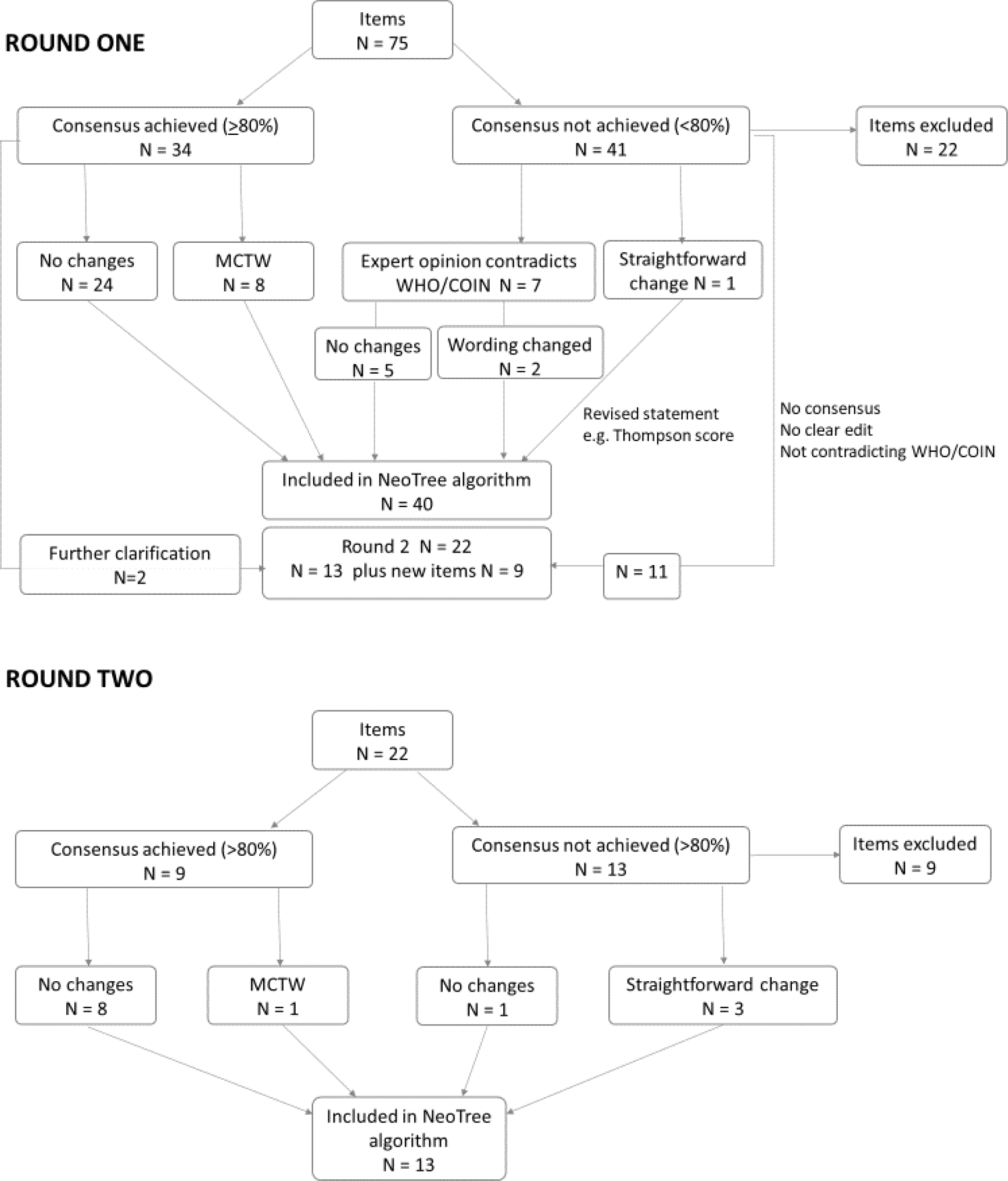
Outcome of algorithm items after round one and round two of the Delphi technique. Abbreviations: COIN = Care of the Infant and Newborn in Malawi; MCTW = minor changes to wording; WHO = World Health Organisation

The expert panel consistently stated that algorithm items must comply with WHO danger signs and COIN guidelines for neonatal sepsis, irrespective of whether the panel agreed with them. For example, experts thought that ‘bulging fontanelle’ was “*subjective; there are non-infectious causes … many babies’ fontanelles bulge when they just cry.”* Another item that did not meet consensus was ‘poor feeding,’ which experts found vague for multiple reasons, including: *“it is too subjective; it depends on how long for … many newborns do not feed well on the first day of life*.” However, experts agreed that poor feeding was a sign of possible sepsis if it was *“a new onset of poor feeding when the infant had previously been feeding well.”* This item was changed to ‘new onset of poor feeding’ for the final algorithm.

Two items that were included because they are part of the COIN guidelines highlighted inconsistencies with WHO guidelines. For example, COIN uses a temperature of more than 37.5°C as a fever for a newborn, whilst WHO and most experts use more than 38°C. Therefore, 37.5°C was included for the Malawian digital platform, but 38°C will be used for other countries. Other items where a difference between the recommendations and the guidelines occurred were antibiotic choice and duration for neonatal sepsis.

Modifications usually involved adopting the language utilized by WHO or COIN, but there were items that experts felt needed clarifying. For example, experts felt that *“twitching or abnormal movements”* needed to be added to the WHO term ‘convulsions’ because seizures in a neonate can be very subtle. Certain items that that could not be revised easily were submitted for the second round according to feedback from the expert panel. For example, experts disagreed that ‘very/extremely premature (<32 weeks gestation)’ was a major risk factor for sepsis if *“the baby was delivered as a clean cold caesarean section for maternal reasons and the mother was not in labour.”* Eighty percent of experts highlighted that the algorithm should include weight to guide gestation because “*gestation is often unknown*” and “*you are relying on [the] Ballard score which has plus or minus two weeks accuracy.”* Similar opinions regarding method of delivery and the importance of birth weight were expressed for ‘slightly premature (32-36 weeks gestation).’ Both gestational age brackets were submitted into the second round as risk factors for sepsis after modifying the items to include WHO weight parameters to guide gestation. Other items that did not gain consensus and were submitted for the second round included items that experts felt needed further clarification. ‘Born before arrival’ as a minor risk factor for sepsis was clarified to the experts that this meant the baby was born en-route to the hospital (either in a vehicle or on the roadside, both being considered dirty environments in Malawi). A ‘neonate admitted with or history of a fever’ as a minor risk factor for sepsis was changed to ‘mother reports a non-measured fever’ in the second round. Lastly, because experts considered the term ‘birth injury’ unclear, we asked them in round two to define what they considered a ‘significant birth injury.’

### Key findings by algorithm

The first important finding was that the ‘major’ or ‘minor’ algorithmic weighting system (where one major risk factor for sepsis is equivalent to two minors) used to diagnose neonatal sepsis was near consensus (77%) but did not meet the 80% threshold. Experts called for further evidence before adopting this system: *“It is a difficult thing to do…you need to work out how specific and sensitive the app is by looking at blood cultures.”* Two experts suggested using a risk score calculator, and another two experts highlighted that WHO only uses danger signs. This weighting system was subsequently removed from the algorithm (Figure 2).

**Figure 2.**
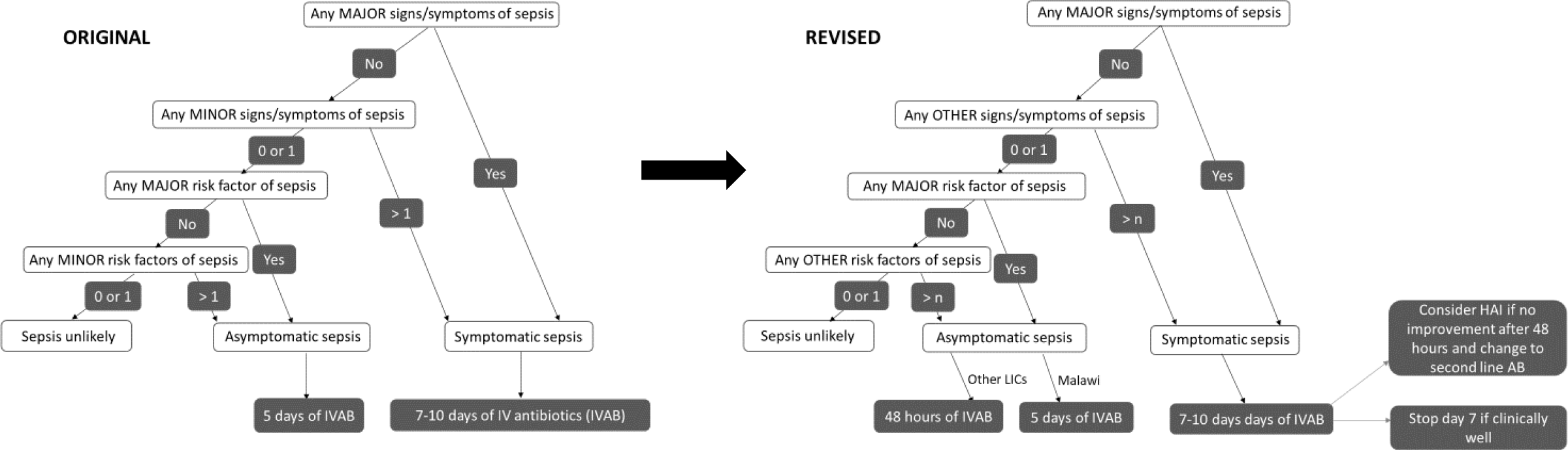
Modification of the neonatal sepsis algorithm as a result of the Delphi technique. Abbreviations: AB = antibiotics; HAI = hospital-acquired infection; IVAB = intravenous antibiotics; LICs = low-income countries; > n = number of risk factors to be determined

The second significant algorithmic finding was on HIE. An academic expert in neonatal encephalopathy discouraged the use of the term ‘birth asphyxia,’ a term used by Malawian HCWs and therefore incorporated into the original algorithm.

*You really must not call it birth asphyxia because birth asphyxia means failing to breathe at birth and what you are talking about is encephalopathy*.

Additional feedback on the algorithm focused on the combination of risk factors or clinical signs and symptoms to consider or diagnose HIE. Experts cited a lack of evidence for using risk factors to diagnose birth asphyxia and that the application should only be using clinical signs and symptoms.

*Birth asphyxia is not about risk factors. If you have encephalopathy it is a clinical diagnosis, and it is irrelevant what your risk factors are*.

Experts recommended utilizing a validated encephalopathy score,[19, 20] which was incorporated into the HIE algorithm. The risk factors that met consensus will be used as prompts to perform the Thompson score, which uses clinical signs and symptoms exclusively to diagnose HIE.

Third, for the respiratory algorithm, experts highlighted that “*it is hard to make an accurate diagnosis of a respiratory condition without investigations*.” Therefore, the algorithm should focus instead on the management of respiratory distress. All respiratory conditions (respiratory distress syndrome, meconium aspiration, congenital pneumonia and transient tachypnoea of the newborn (TTN)) now fall under the umbrella diagnosis of respiratory distress of the newborn within the algorithm. For teaching purposes, the four respiratory conditions will be included as ‘diagnoses to consider’ in the management.

Finally, for the hypothermia algorithm, experts commented that first-line treatment for all newborns be skin-to-skin care including those who were severely hypothermic (<32°C) unless they showed any signs or symptoms of being unstable. Additionally, experts did not think it was realistic to review a newborn every 15-30 minutes when hypothermic.

### Round Two

Ten (71%) experts completed round two, seven electronically and three by telephone interview. Four experts dropped out (three from HICs, one from LIC); three did not respond to email reminders and one expert was unable to meet the completion deadline. Nine items (41%) reached consensus (Figure 1). These items were either (1) included, unmodified (36%) or (2) included, modified in the revised algorithm (5%). Items that did not reach consensus (59%) were either (1) excluded (41%) or (2) included, modified according to WHO guidelines or expert suggestion (18%).

In round one experts indicated that hypothermia was a major sign of sepsis and should be included in the sepsis algorithm if persistent. In round two we clarified that the application is to be used at the time of admission onto the neonatal unit, at which point the HCW will only have one temperature reading. Experts in round two disagreed that a single temperature reading of <35.5_JC was a sign of sepsis and felt that it would more likely be due to environmental hypothermia, a common problem in LICs. Additionally, in round two it was established that experts were much more concerned with extremely premature <1500g neonates (88% consensus) compared to slightly premature 1500-2500g neonates (62% consensus) being at risk for neonatal sepsis. Central cyanosis was an addition to the second round as an expert suggestion to include all WHO danger signs; despite missing consensus (with 78% agreement), it was ultimately included in the final sepsis algorithm to comply with WHO guidelines for danger signs. All of the respiratory items marginally missed consensus. All three items were still included in the final teaching algorithms with revisions in line with WHO diagnostic criteria. Modifications to the algorithms can be found in online supplementary file 2.

## DISCUSSION

We report the use of a modified Delphi technique to review digital clinical pathway algorithms for four neonatal conditions managed by HCWs in LICs. Approximately two thirds (62%) of the original algorithm items were ultimately included for use in the NeoTree digital platform based on consensus expert opinion and national/international guidelines. The NeoTree team revised the algorithms based on this feedback. Expert discussion emphasised gaps in evidence in neonatal care in LICs, highlighting areas for future research.

Each algorithm had components that triggered debate amongst the experts. For neonatal sepsis, three points were discussed. First, experts called for further evidence before adopting a ‘major’ and ‘minor’ algorithmic weighting system to diagnose neonatal sepsis. In response the NeoTree research team are conducting a study in Zimbabwe and Malawi looking at which clinical indicators are predictors of positive blood cultures. Second, there was disparity in opinion regarding whether to give prophylactic antibiotics and the duration of antibiotics for newborns with risk factors for sepsis who remain clinically well without any supporting investigations (NeoTree’s equivalence to asymptomatic sepsis). The WHO recommendation to administer prophylactic antibiotics for a neonate with maternal risk factors for sepsis only is considered weak with very low-quality evidence.[21] Despite reaching a consensus on particular risk factors (prolonged rupture of membranes, maternal fever), experts also highlighted the evidence base as weak. In terms of duration of treatment for asymptomatic sepsis, while expert opinion varied, the Malawian guidelines recommend a five-day course[11] whilst WHO recommends two days.[16] The NeoTree algorithms will therefore keep to local and international recommendations, but the NeoTree team will feed back to the Malawian COIN expert panel that consensus suggested five days is too long to treat newborns with sepsis risk factors only. Third, experts disputed the treatment choice and duration for symptomatic neonatal sepsis; incidentally, WHO recommendations lack strong evidence or efficacy.[22]

For the HIE algorithm, the Thompson score was preferred because it is simpler to perform, less time consuming and better at predicting poor outcomes in moderate and severe HIE during the first hours of life compared to the Sarnat score at 24 hours.[19] The NeoTree research team suspected that measures such as examining for posturing and Moro reflex would be relatively complicated for frontline HCWs with minimal training to assess. However, neonatal experts’ experience and previous studies in LICs[23] assured the team that the score is relatively straightforward to teach.

Several points of discussion also centred on the respiratory algorithms. First, experts noted that even with investigations in HICs respiratory conditions may be difficult to diagnose.[24] Second, despite experts’ concerns about antibiotic overprescribing in LICs and the need to differentiate TTN from other respiratory conditions, they did not think this was currently feasible in LICs due to limitations in HCW capacity, resources and knowledge. Thus, experts agreed that all neonates with signs of respiratory distress should have respiratory support and antibiotics. A recent study justified the use of antibiotics for tachypnoea alone in a neonate in a resource-limited setting.[25] Third, experts recommended performing chest x-rays (if available) only if imaging would change management (e.g. a longer course of antibiotics for congenital pneumonia) or if the neonate was deteriorating.

With the proliferation of clinical digital platforms in HICs and LICs, there is growing concern with the quality and safety standards of their clinical guidance. Countries and organisations (including WHO) are now taking measures to ensure application developers fulfil a strict set of criteria to protect patients.[26] While the Delphi technique can establish expert consensus, it may also strengthen the safety and quality standards of clinical algorithms. This technique has been widely used in developing paper-based neonatal clinical guidelines in HICs and LICs.[27-29] There are also studies that have used the Delphi technique to develop items used in m-health tools.[30-32] Our study is unique in the application of this technique to develop algorithms on a digital platform specific to neonatal care in low-resource settings.

This study has several limitations. The choice of using a modified two-step Delphi process meant that a final face-to-face meeting was not possible, which may have prevented some exchange of important information to clarify differences in expert opinion. However, this method allowed for the contributions of geographically dispersed experts, maintained their anonymity and prevented them from conforming to other experts. The recruitment of more experts from HICs (64%) compared to LICs (36%), despite originally inviting equal numbers to participate, could have contributed to expert panel bias. Dropouts from the first to the second round could have affected the consensus level and contributed to attrition bias.

Some factors may have contributed to selection bias. The Delphi process is time intensive, which could have meant that those clinicians who are busier with perhaps even more clinical expertise or those with limited internet access (mainly LICs) could not participate. Additionally, offering a financial incentive might have obtained a more equal representation of experts. Another drawback of the Delphi being a labour-intensive process was that a year elapsed between the two rounds. Experts may have forgotten the algorithms and items from the first round in the second round if they did not read the summary of results or refresh their knowledge of the algorithms. Experts reported that they found the layout of the second questionnaire confusing; a re-design contributed to delays.

This study used the Delphi technique to refine four clinical decision algorithms in a neonatal digital platform designed for HCWs in LICs to standardise and improve the quality of newborn care. The key to implementing the NeoTree algorithms in other LICs will be to demonstrate that clinical algorithms in a digital application versus paper-based guidelines can aid HCWs in making faster, more accurate diagnoses and provide better, more cost-effective treatment that will ultimately improve the quality of newborn care and reduce mortality. This will require a large-scale clinical-trial evaluation. Ultimately, with consensus opinion shaping the algorithms of this digital platform, accurate data capture, immediate clinical assessment and optimal medical care may be achieved to improve neonatal outcomes.

## Data Availability

All data relevant to the study are included in the article, uploaded as supplementary information or available upon request from the corresponding author.

## ACKNOWLEDGMENTS

We thank all the participants of the two rounds of Delphi surveys for their invaluable contributions to revise the clinical algorithms and highlight open questions. We would also like to thank Dr. Brian Corden and Dr. Alice Myers for their reviews of the manuscript.

## CONTRIBUTORS

ME, CC and MH conceived the study, generated the methodology and designed the questionnaires. ME conducted the interviews and analysed the data. ME and MC produced the first draft and contributed equally to this manuscript. CC, FF and MH provided edits and comments to the draft. All authors reviewed and approved the final version of the manuscript.

## ETHICS APPROVAL

Ethics approval was not required for this study according to the University College of London Research Ethics Committee.

## PROVENANCE AND PEER REVIEW

Not commissioned; externally peer reviewed.

## PATIENT CONSENT FOR PUBLICATION

Not required.

## ORCID IDs

Mari Evans https://orcid.org/0000-0001-5113-3629 Mark Corden https://orcid.org/0000-0001-6146-1634 Caroline Crehan https://orcid.org/0000-0002-3655-6954

Felicity Fitzgerald https://orcid.org/0000-0001-9594-3228 Michelle Heys https://orcid.org/0000-0002-1458-505X

